# Are News Media Organizations Protecting Youth Mental Health? A Content Analysis of the Top 4 U.S. News Organizations in the on TikTok

**DOI:** 10.64898/2026.01.21.26344595

**Authors:** Caroline A.P. Helsen, McKenna F. Parnes, Dara E. Gleeson, Hannah Scott, Victoria McClare, Rhayna Poulin, Sarah R. Lowe

## Abstract

In 2021, the U.S. Surgeon General issued an advisory to address the youth mental health crisis that included actions media organizations could take regarding their coverage of mental illness and traumatic events including natural disasters, pandemics, and mass violence. Although research indicates that both news and social media are associated with worse adolescent mental health outcomes, it is unknown whether news outlets adhere to the U.S. Surgeon General’s recommendations on social media. This study aimed to describe how news media organizations adhered to federal recommendations in their reporting of mental health or illness and collective trauma events on the TikTok social media platform. We conducted a directed content analysis of TikTok videos posted by four leading U.S. news organizations from December 2021 to December 2022 Videos covering relevant topics were coded based on their adherence to advisory guidelines. Of 5,344 TikTok videos posted during the timeframe, 950 (17.8%) covered news related to mental health or illness, climate change or weather-related disasters, mass shootings, and the COVID-19 pandemic. Content had a median adherence rate of 19.5% (range: 0.0-83.9%); however, substantial variability was observed across guidelines, news organizations, and topics. The two recommendations with the highest adherence rate were “fact-based reporting” and “no language that shocks, provokes, or creates a sense of panic” (40.0-76.1%). The recommendations with the lowest adherence rate (0.0-8.0%) were “include ways the public can make a positive difference” and “include content warnings on distressing content.” Findings can inform news media efforts to develop content that protects youth mental health and interventions that support youth news consumption on social media.

**Author Summary:** Young people across the United States (U.S.) turn to social media for news about current events. Unfortunately, many social media posts from news media organizations amplify distressing events, and seeing this content has negative consequences for youth mental health. Recognizing the potential ways in which social media can affect youth mental health, the U.S. Surgeon General issued an advisory to address the youth mental health crisis that included actions news media organizations could take when sharing content of mental illness and traumatic events. Here, we use content analysis methods to analyze whether news media organizations followed the federal recommendations when covering mental illness and collective trauma events on TikTok. Content that covered these topics had a median adherence rate to the Surgeon General recommendations of 19.5% (range: 0.0-83.9%); however, substantial variability was observed across guidelines, news organizations, and topics. Our study offers new insights public health leaders can use to inform efforts to promote media content that protects youth mental health.

## Introduction

From 2011 to 2021, adolescents in the United States experienced significantly worse mental health, with a 14.0% increase in students experiencing persistent feelings of sadness or hopelessness and a 6.0% increase in students seriously considering suicide (1). Recognizing the significance of this trend, the U.S. Surgeon General’s Office issued an advisory on December 7, 2021 that called for action to address the youth mental health crisis (2).

The advisory, titled “Protecting Youth Mental Health,” puts forth a set of recommendations for whole-of-society efforts to improve youth mental health (3). One set of recommendations in the advisory highlights actions media organizations can take to protect viewers’ mental health including ways to report negative news and stories about mental health and illness. The advisory mentioned negative news coverage of natural disasters, pandemics, and mass violence, also known as collective trauma events. Collective trauma events can be characterized as long-term with an unclear endpoint and a broad population-level impact (4). Coverage of these events can contribute to viewers’ psychological distress, (5). For example, a longitudinal study found that media exposure about the 2013 Boston Marathon bombing predicted post-traumatic stress symptoms. After the 2016 Orlando Pulse nightclub mass shooting, subsequent worry about future traumatic events predicted an increase in media consumption and acute stress (6). Regarding coverage of mental health and illness, false or misleading stories can perpetuate stigma (7), which can lead to delays in seeking and receiving treatment (8).

The concern regarding news media and mental health is made worse by the high percentage of youth who consume news on social media, a frequently cited factor affecting youth mental health (9). Ninety-three percent of youth, specifically Generation Z born between 1997 and 2012, consume news content on social media weekly (10). Top Gen Z social media platforms for news include: Instagram, TikTok, and Snapchat (11). While political and national security concerns regarding TikTok call into question the app’s future in the U.S. (12), it is clear the app is growing in popularity. A Google Executive acknowledged the uptick in Gen Z using the app as a search engine instead of Google Search (13) and the Pew Research Center reported an increasing trend in American users consuming news on TikTok, whereas apps like Snapchat, YouTube, and Instagram are seeing declining or stagnant trends (14).

Existing research examines the negative impact on mental health as it relates to each domain separately: news media or social media. However, there is a gap in research that characterizes social media content produced by news organizations as it relates to youth mental health. One systematic review analyzed 25 studies that primarily used a content analysis approach to describe TikTok content about public and mental health topics such as eating disorders and the COVID-19 pandemic; however, the content sources were public health accounts, charities, and personal accounts (15). Studies about news media on TikTok focused on how companies are adapting to TikTok culture through use of text on screen, filters, stickers, original audio clips, and funny challenges (16). To the authors’ knowledge, no published studies have examined how news media organization content on TikTok may impact mental health and there have been no attempts to examine whether news outlets adhere to the U.S. Surgeon General’s recommendations on or off social media.

This study aims to describe how news media organizations adhere to federal recommendations to protect youth mental health while reporting stories on mental health or illness and collective trauma events, including climate change or weather-related disasters, mass shootings, and the COVID-19 pandemic.

## Methods

The study used a directed content analysis method to examine news media coverage of collective trauma events and mental health. The content analysis was directed by the U.S. Surgeon General advisory recommendations for media organizations to protect youth mental health. TikTok was selected due to the rising popularity of consuming news on the app and the high percentage of youth who consume news on social media generally.

### Inclusion Criteria

Collective trauma events included in the analysis were: weather-related disasters, climate change (17), the COVID-19 pandemic (3), and mass shootings (18). Other content included in the analysis were any videos mentioning mental health or mental illness topics, such as exercising to improve your mental health, youth mental health in the face of gun violence, celebrities diagnosed with mental illness, substance use disorder, and antipsychotic medication.

We selected news media organizations based on their follower count. Of the top 50 news sites in the United States (19), only 32 are on TikTok. Excluding foreign-based news organizations, such as The Daily Mail, the top four news organizations on TikTok based on follower count are: ABC News (4.6 million), NBC News (3.8 million), CBS News (3.0 million), and Daily Wire (2.7 million). ABC News, NBC News, and CBS News represent the broadcast networks in the United States. Fox News, a cable network, is the main news source for nearly half of conservatives (20). While Fox News is not on TikTok, Daily Wire describes itself as right-of-center media (21) and is a popular conservative news source (22).

The study period was December 7, 2021, when the Surgeon General advisory was published, through to December 6, 2022. This year-long time frame allowed for greater content volume and variability resulting in a more comprehensive analysis. Patient consent statement is not applicable or needed for this study.

### Data Collection and Processing

A Qualtrics survey was used to code all videos from ABC News, NBC News, CBS News, and Daily Wire posted on TikTok between December 7, 2021 to December 6, 2022. Data collected for all videos included: URL link to the post, date of post, number of likes, number of comments, main topic using IPTC media topic categories (23), and subtopic. If the subtopic did not fit the study’s inclusion criteria, the survey terminated. Table 1 lists the Surgeon General recommendations for mental health and illness content and the corresponding coding survey question.

**Table 1.**
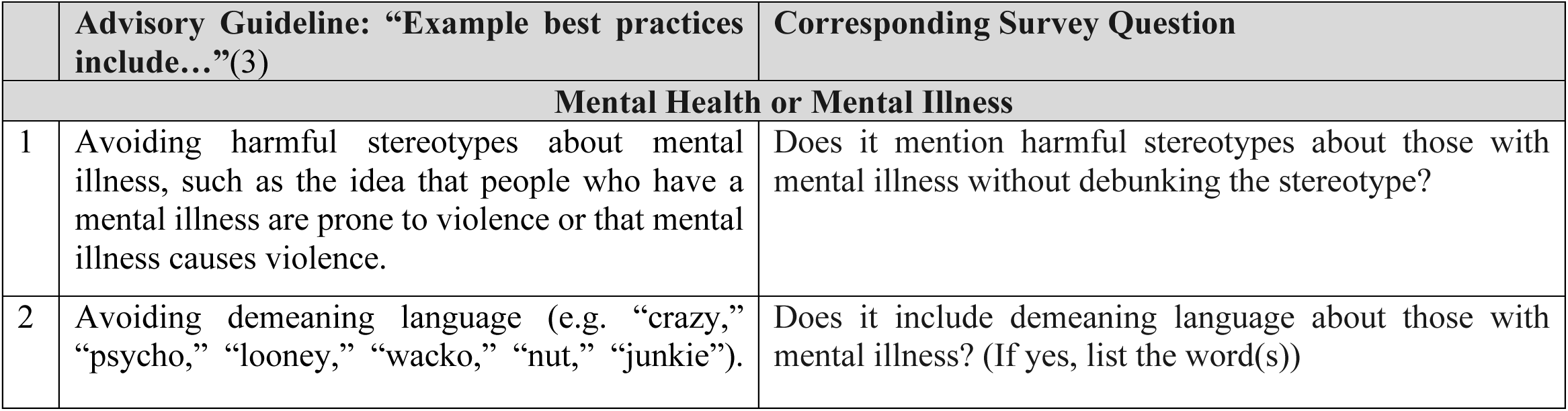
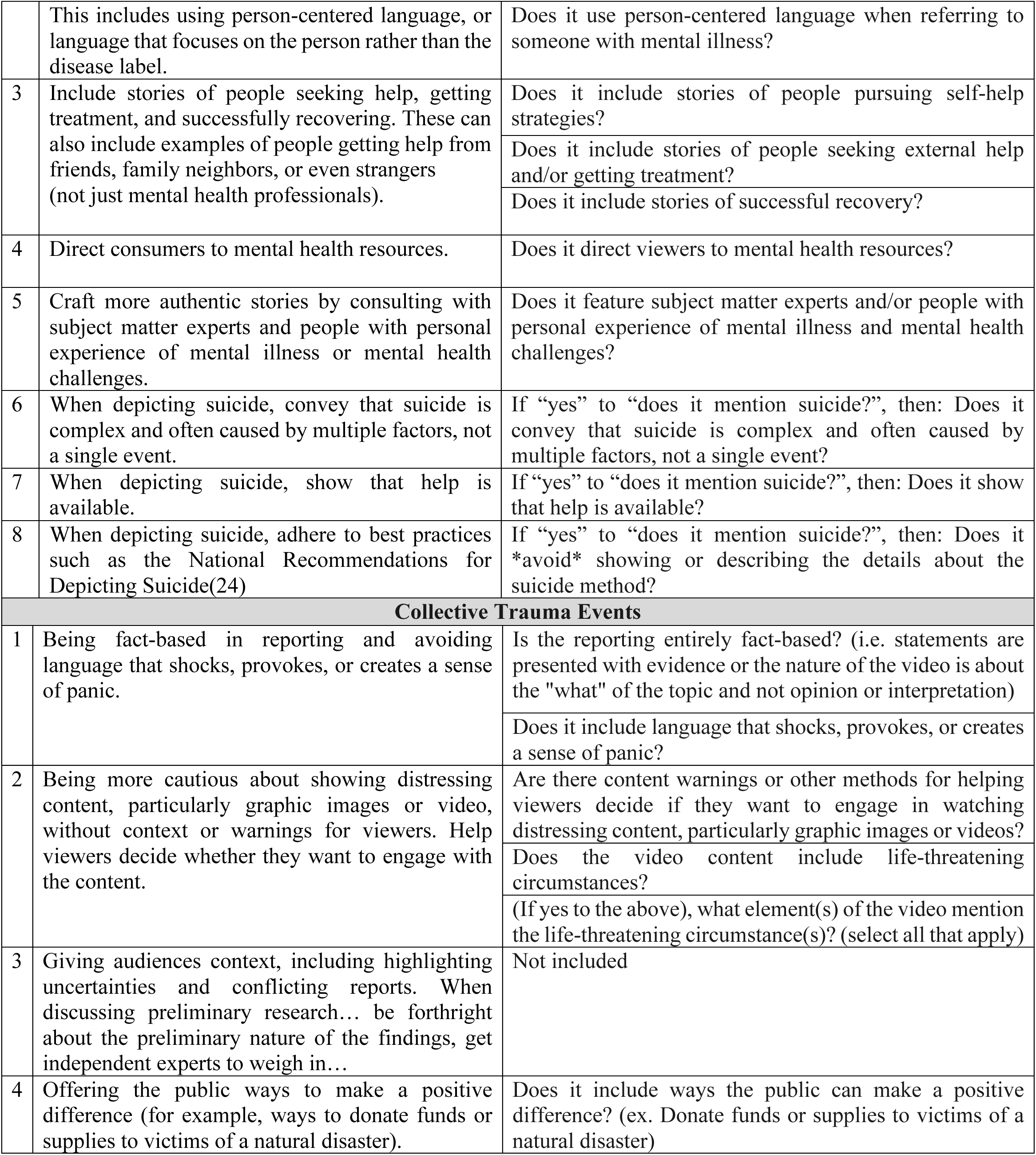
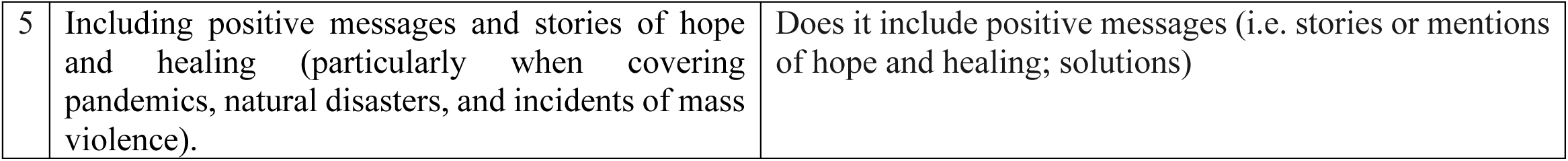
Survey questions for videos meeting inclusion criteria for mental health or mental illness and collective trauma events.

Table 2 similarly lists the recommendations and corresponding questions, but for collective trauma content. As shown in the Additional Information column, some recommendations provided only broad guidance and needed further definition. For example, the advisory did not define language that shocks, provokes, or creates a sense of panic, so this study used a narrow definition to include content that: says “shock*,” says “panic*,” has a hopeless or doomsday message, indicates you are in danger, or makes a claim or accusation about a group of people. . While the advisory only mentions individual-level action such as donating supplies during a disaster, it is also important for news media to highlight community- and structural-level action. We added and collected data for a recommendation to include actions that address the issue at a level beyond the individual.

**Table 2.**
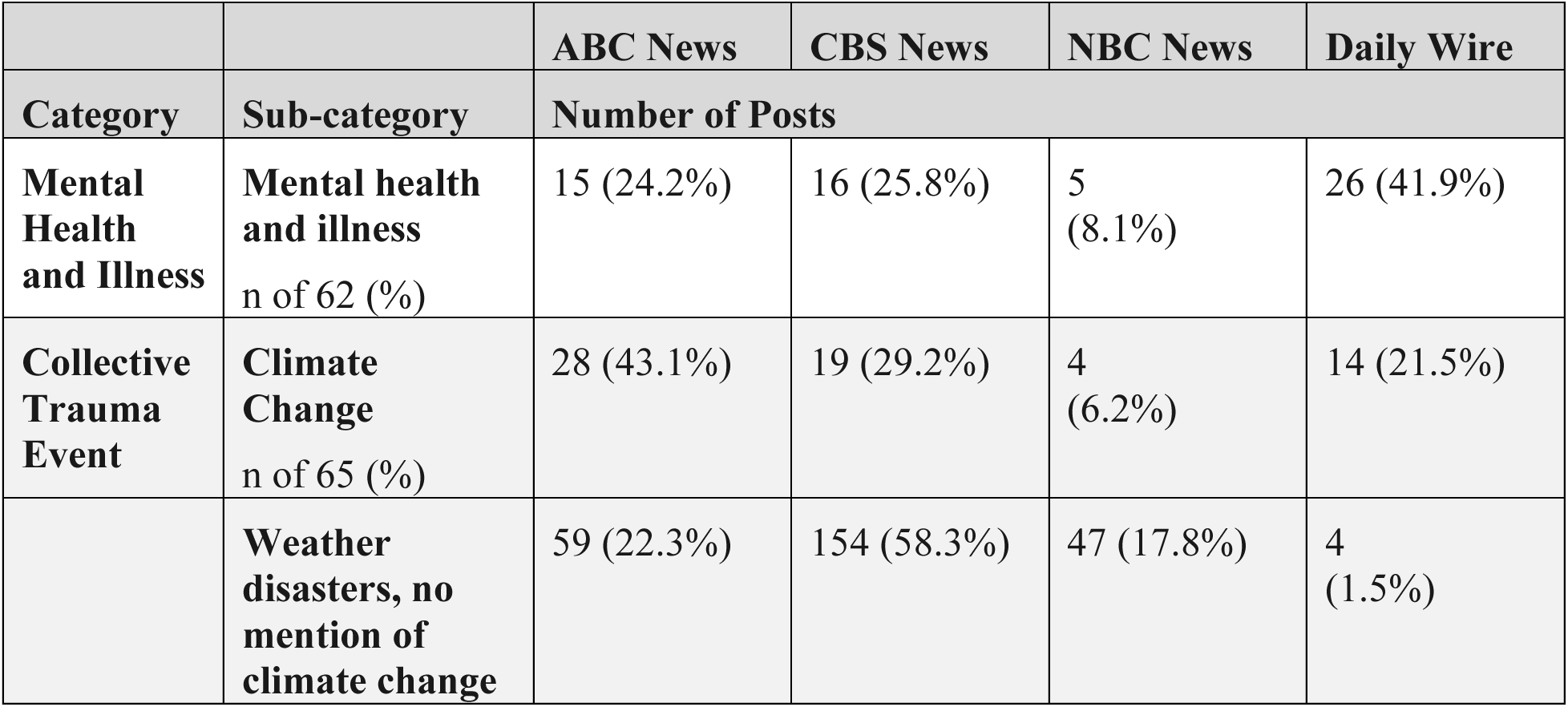
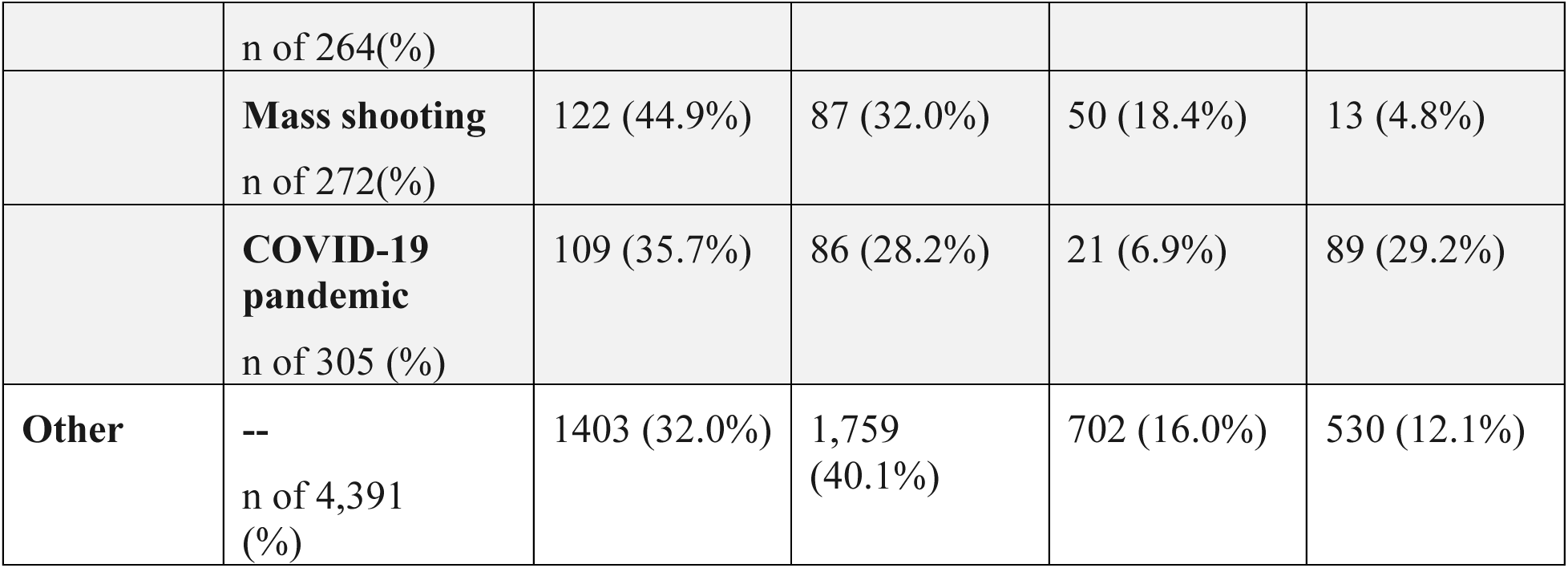
Characteristics of all videos published December 2021 to December 2022 by News Media Organization.

|  |  | ABC News | CBS News | NBC News | Daily Wire |
| --- | --- | --- | --- | --- | --- |
| Category | Sub-category | Number of Posts |  |  |  |
| **Mental Health and Illness** | **Mental health and illness**<br>n of 62 (%) | 15 (24.2%) | 16 (25.8%) | 5 (8.1%) | 26 (41.9%) |
| **Collective Trauma Event** | **Climate Change**<br>n of 65 (%) | 28 (43.1%) | 19 (29.2%) | 4 (6.2%) | 14 (21.5%) |
|  | **Weather disasters, no mention of climate change** n of 264(%) | 59 (22.3%) | 154 (58.3%) | 47 (17.8%) | 4 (1.5%) |
|  | **Mass shooting**<br>n of 272(%) | 122 (44.9%) | 87 (32.0%) | 50 (18.4%) | 13 (4.8%) |
|  | **COVID-19 pandemic**<br>n of 305 (%) | 109 (35.7%) | 86 (28.2%) | 21 (6.9%) | 89 (29.2%) |
| **Other** | --<br>n of 4,391 (%) | 1403 (32.0%) | 1,759 (40.1%) | 702 (16.0%) | 530 (12.1%) |

The coding team consisted of four Master of Public Health candidates and one Bachelor of Science candidate, supervised by one Associate Professor thesis advisor. We held a meeting to discuss inclusion criteria and the Qualtrics survey. All coders analyzed the same set of 24 videos and held a subsequent meeting to review results and discuss clarifications for future coding. The team achieved 90.3% agreement on coding not including irrefutable survey answers such as the date of the post. With a high level of agreement, the team decided to proceed with dividing and coding all videos. To ensure data quality throughout the coding process, the team discussed any remaining questions on a daily basis via email and during two additional meetings before coding the videos in question.

Further data integrity measures were taken during the data processing phase. We double coded all mental health and illness videos and reconciled any discrepancies. Of the collective trauma event content, 5.0% (n=45) were randomly sampled and double coded. The agreement rate was 94.4%.

### Quantitative and Qualitative Content Analysis

Descriptive statistics, including medians and ranges for count variables, and frequencies and percentages for dichotomous variables, were calculated. To operationalize the Surgeon General’s guidelines and provide tailored recommendations for improvements, data were disaggregated by news media organization.

For qualitative analysis, the process was organized by subtopic: (1) mental health or illness (2) climate change or weather-related disasters with or without mentions of climate change (3) mass shootings and (4) the COVID-19 pandemic. Team members inductively coded videos and included a brief summary of each video in the survey. Inductive codes and summaries were analyzed by the first author for themes.

## Results

ABC News, CBS News, NBC News, and Daily Wire published a total of 5,344 videos from December 7, 2021-December 6, 2022. Of those videos, 950 (17.8%) covered news related to mental health or illness, climate change or weather-related disasters, mass shootings, and the COVID-19 pandemic. Across all exposure types, results show a median adherence rate to the U.S. Surgeon General recommendations to protect youth mental health of 19.5% (range=0.0-83.9%). The recommendation with the highest overall adherence rate across all four platforms (83.9%) was: do not include harmful stereotypes about mental illness. Two recommendations had a 0.0% adherence rate: include content warnings on distressing pandemic content and include ways the public can make a positive difference in weather-related disaster content with no mention of climate change. ABC News had a median adherence rate of 34.6% (range=0.0-95.1%), followed by CBS News at 14.3% (range=0.0-100.0%), NBC News at 9.8% (range=0.0-100.0%), and the Daily Wire at 7.7% (range=0.0-75.0%).

### Overall Content Characteristics

Only 1.2% of all videos were about mental health or illness. ABC News, NBC News, and Daily Wire had 2 to 8 times more posts about mental illness than mental health, whereas CBS News had an equal number of videos on both topics. News coverage of collective trauma events amounted to 17.0% of all published videos. The topic with the most coverage was the pandemic (5.6%), followed by mass shootings (4.9%), weather-related disasters with no mention of climate change (4.9%), and climate change (1.2%).

### Content that Covered Both Mental Health or Illness Stories and a Collective Trauma Event

A total of 18 videos fell into both categories of mental health or illness stories and collective trauma event. Ten (55.6%) were about mass shootings and eight (44.4%) were about the COVID-19 pandemic. Table 3 shows the number and percentage of videos by news media organizations on collective trauma events that met each Surgeon General recommendation from 2021.

**Table 3.**
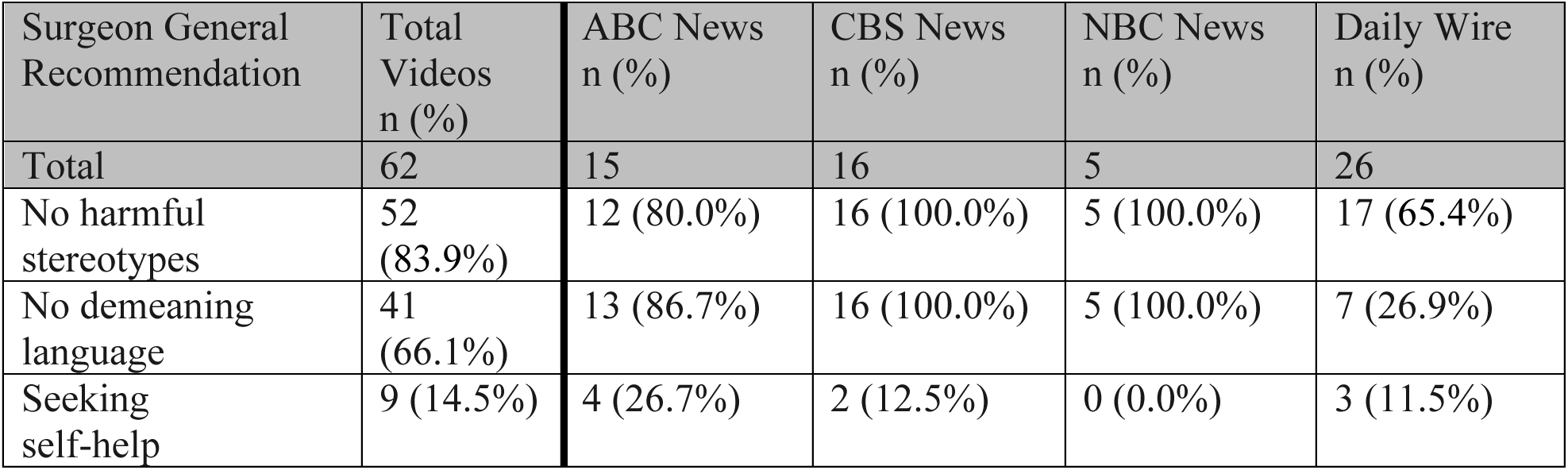
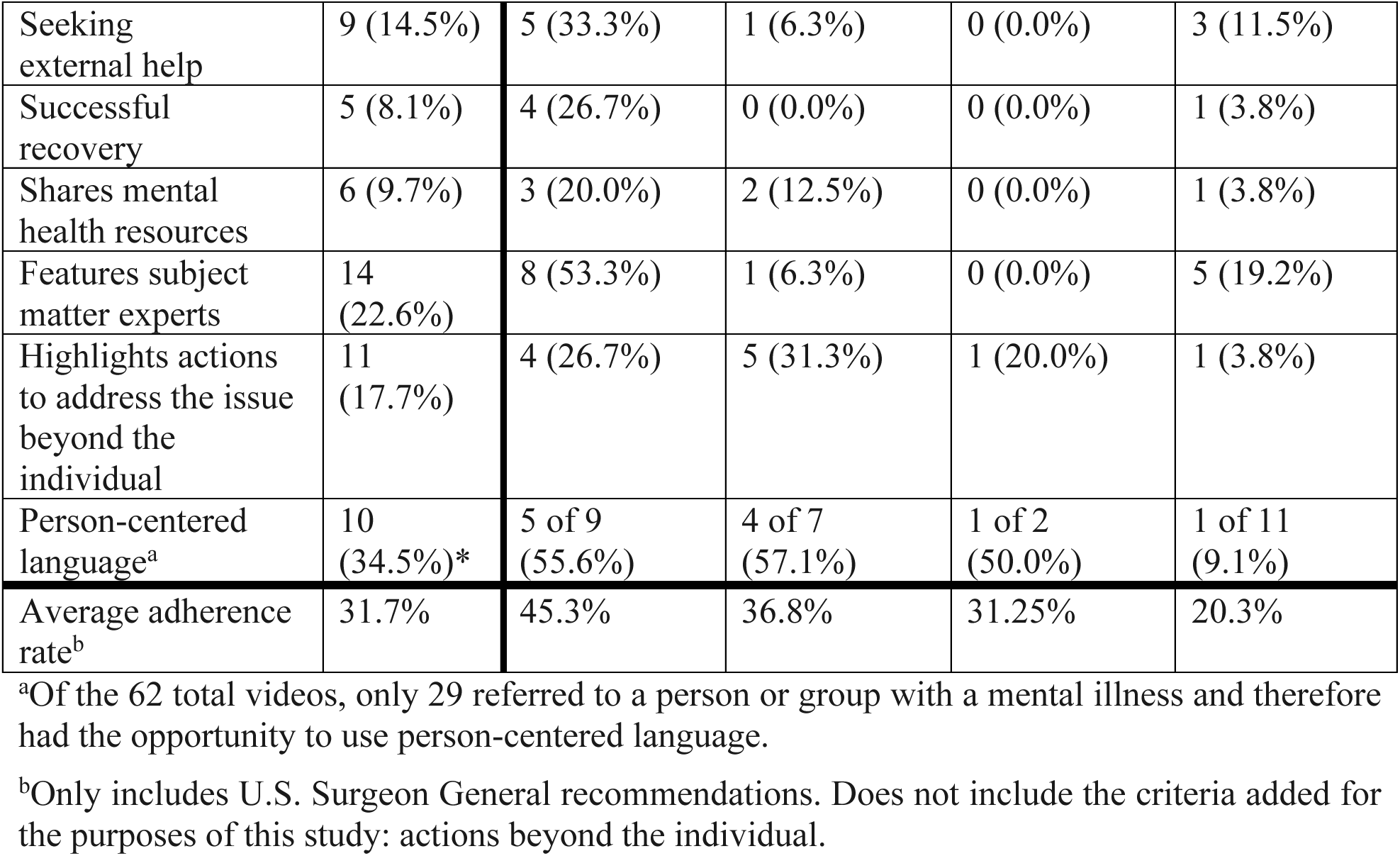
Mental health and illness videos that follow 2021 Surgeon General recommendations by news media organization.

| Surgeon General Recommendation | Total Videos n (%) | ABC News n (%) | CBS News n (%) | NBC News n (%) | Daily Wire n (%) |
| --- | --- | --- | --- | --- | --- |
| Total | 62 | 15 | 16 | 5 | 26 |
| No harmful stereotypes | 52 (83.9%) | 12 (80.0%) | 16 (100.0%) | 5 (100.0%) | 17 (65.4%) |
| No demeaning language | 41 (66.1%) | 13 (86.7%) | 16 (100.0%) | 5 (100.0%) | 7 (26.9%) |
| Seeking self-help | 9 (14.5%) | 4 (26.7%) | 2 (12.5%) | 0 (0.0%) | 3 (11.5%) |
| Seeking external help | 9 (14.5%) | 5 (33.3%) | 1 (6.3%) | 0 (0.0%) | 3 (11.5%) |
| Successful recovery | 5 (8.1%) | 4 (26.7%) | 0 (0.0%) | 0 (0.0%) | 1 (3.8%) |
| Shares mental health resources | 6 (9.7%) | 3 (20.0%) | 2 (12.5%) | 0 (0.0%) | 1 (3.8%) |
| Features subject matter experts | 14 (22.6%) | 8 (53.3%) | 1 (6.3%) | 0 (0.0%) | 5 (19.2%) |
| Highlights actions to address the issue beyond the individual | 11 (17.7%) | 4 (26.7%) | 5 (31.3%) | 1 (20.0%) | 1 (3.8%) |
| Person-centered language<sup>a</sup> | 10 (34.5%)* | 5 of 9 (55.6%) | 4 of 7 (57.1%) | 1 of 2 (50.0%) | 1 of 11 (9.1%) |
| Average adherence rate<sup>b</sup> | 31.7% | 45.3% | 36.8% | 31.25% | 20.3% |

<sup>a</sup>Of the 62 total videos, only 29 referred to a person or group with a mental illness and therefore had the opportunity to use person-centered language.
<sup>b</sup>Only includes U.S. Surgeon General recommendations. Does not include the criteria added for the purposes of this study: actions beyond the individual.

When mental health was mentioned in the context of mass shootings, three videos (16.7%) referred to the mental health of the community, specifically the “psychological toll” on survivors. Four videos (22.2%) referred to the mental health of the Uvalde shooter, calling into question whether he was mentally ill and labeling him as a “deranged shooter,” a “violent psychopath,” and “demented” with “evil in his heart.”

Three videos (16.7%) referred to legislation that addresses mental health in the context of mass shootings; however, two of them only referred to the need to invest in mental health programs generally. They did not specify if the legislation would address the mental health of potential shooters or survivors. There were eight videos (44.4%) that mentioned mental health in the context of the COVID-19 pandemic, half of which were from the Daily Wire. Some content in these videos referred to people who took COVID precautions as “crazy.”

The remaining four videos (22.2%) from ABC News and CBS News mentioned the increase in mental health struggles during the pandemic such as depression and anxiety, with two emphasizing the impact on youth.

### Mental Health and Illness

There were 62 total videos about mental health or illness, including 10 that were also about mass shootings and eight that were also about the pandemic. Table 4 shows the number and percentage of videos by news media organizations on mental health and illness that met each Surgeon General recommendation from 2021. Only two recommendations were met more than 50.0% of the time: do not include harmful stereotypes about mental illness (83.9%) and do not include demeaning language (66.1%). The remaining six recommendations were met in less than 35.0% of videos. Stories of successful recovery were only included in 8.1% of videos, the lowest adherence rate for any of the recommendations surrounding mental health that were examined. The ninth recommendation was to highlight actions to address the issue beyond the individual, with only 11 (17.7%) videos adhering to that recommendation.

**Table 4.**
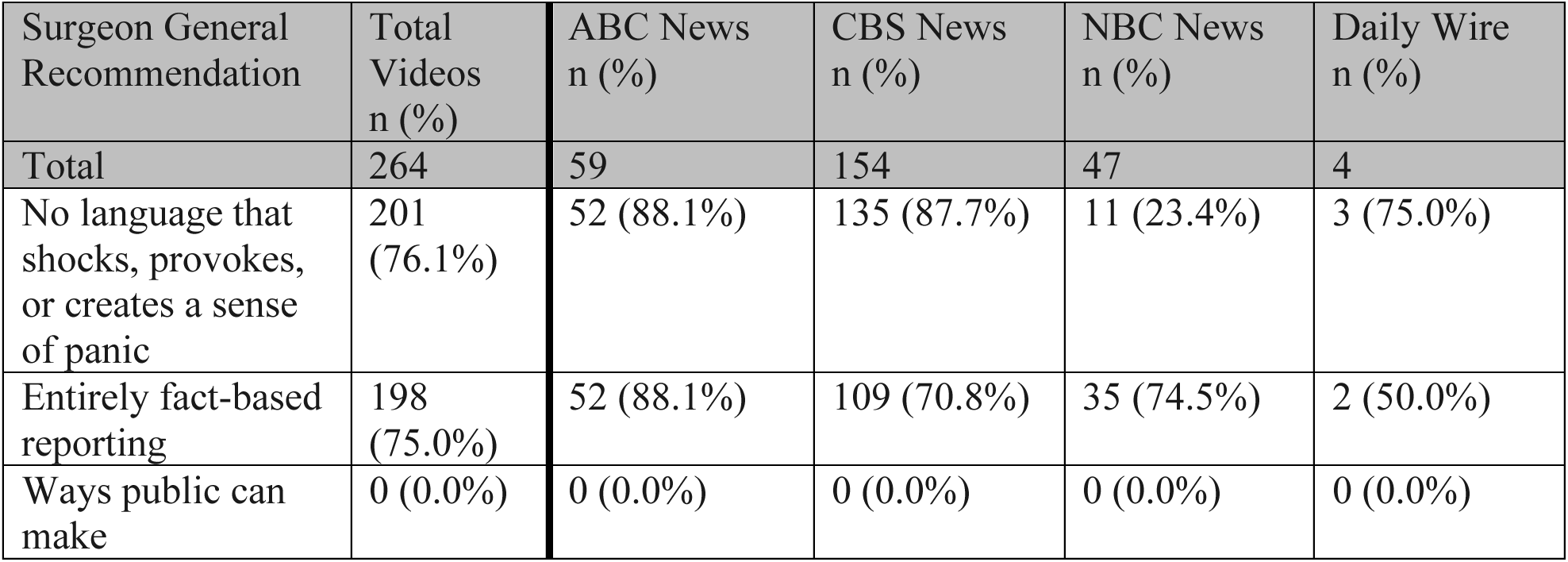
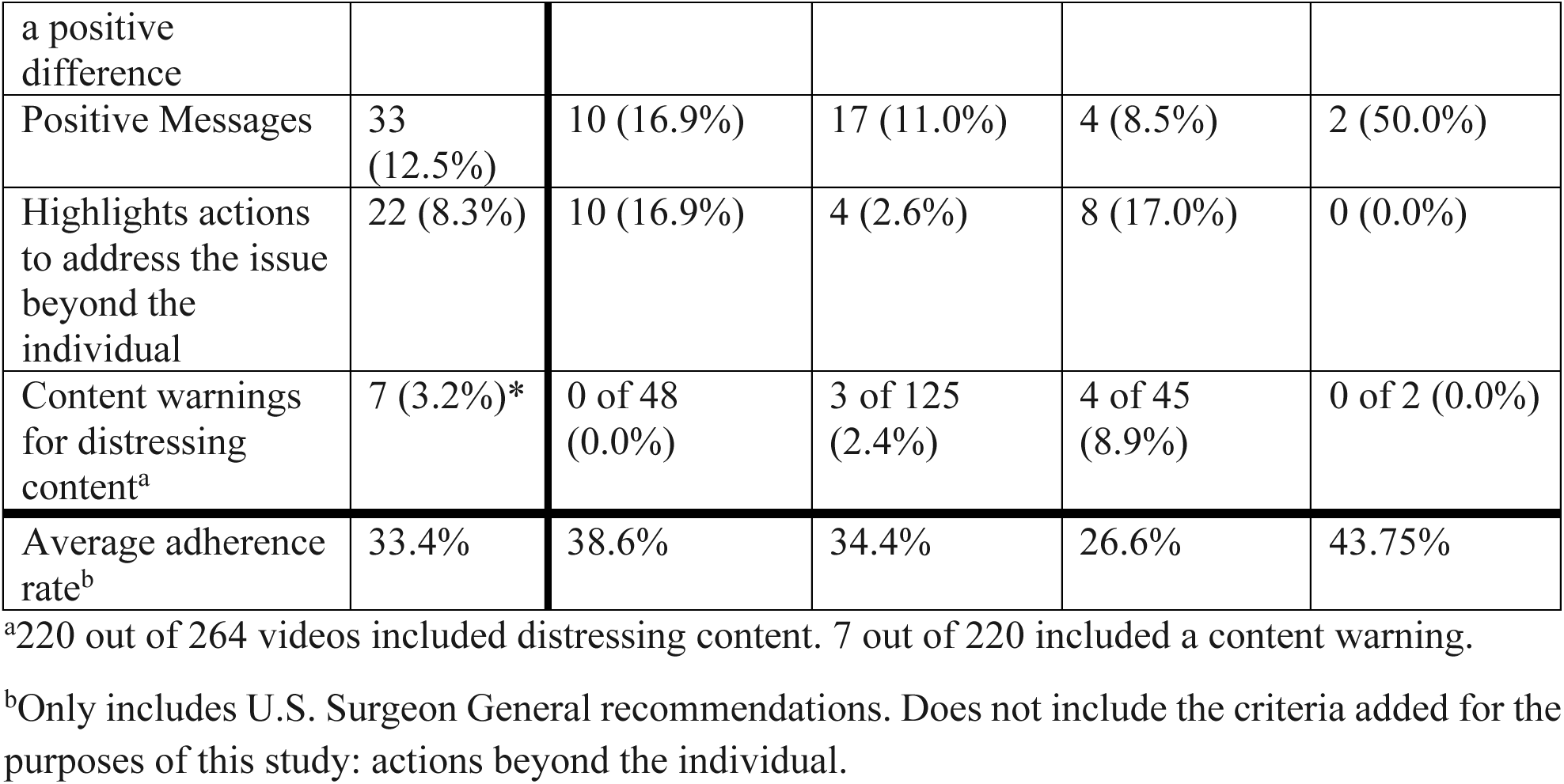
Weather-related disaster videos with no mention of climate change that follow 2021 Surgeon General recommendations by news media organization.

| Surgeon General Recommendation | Total Videos n (%) | ABC News n (%) | CBS News n (%) | NBC News n (%) | Daily Wire n (%) |
| --- | --- | --- | --- | --- | --- |
| Total | 264 | 59 | 154 | 47 | 4 |
| No language that shocks, provokes, or creates a sense of panic | 201 (76.1%) | 52 (88.1%) | 135 (87.7%) | 11 (23.4%) | 3 (75.0%) |
| Entirely fact-based reporting | 198 (75.0%) | 52 (88.1%) | 109 (70.8%) | 35 (74.5%) | 2 (50.0%) |
| Ways public can make | 0 (0.0%) | 0 (0.0%) | 0 (0.0%) | 0 (0.0%) | 0 (0.0%) |
| a positive difference |  |  |  |  |  |
| Positive Messages | 33 (12.5%) | 10 (16.9%) | 17 (11.0%) | 4 (8.5%) | 2 (50.0%) |
| Highlights actions to address the issue beyond the individual | 22 (8.3%) | 10 (16.9%) | 4 (2.6%) | 8 (17.0%) | 0 (0.0%) |
| Content warnings for distressing content<sup>a</sup> | 7 (3.2%)* | 0 of 48 (0.0%) | 3 of 125 (2.4%) | 4 of 45 (8.9%) | 0 of 2 (0.0%) |
| Average adherence rate<sup>b</sup> | 33.4% | 38.6% | 34.4% | 26.6% | 43.75% |

<sup>a</sup>220 out of 264 videos included distressing content. 7 out of 220 included a content warning.
<sup>b</sup>Only includes U.S. Surgeon General recommendations. Does not include the criteria added for the purposes of this study: actions beyond the individual.

Of the four organizations, Daily Wire had the lowest adherence rate to the recommendation of not including harmful stereotypes (65.4%) and not using demeaning language (26.9%) about mental illness. ABC News most consistently met recommendations; however, some were met as low as 20.0% of the time.

Eighteen videos (29.0%) featured stories of people seeking help, with half portraying self-help and half external help. Self-help strategies shared included: sharing your story to connect, cope, and raise awareness; exercise; finding inner strength; reading self-help books; sleeping; “putting yourself first;” and walking in nature. External help or treatment methods shared included: seeking therapy, emotional support animals, connecting with religious leaders, attending AA meetings, attending rehab, and using prescription medication. Seven out of 18 (38.9%) featured celebrities. The criteria news media organizations adhered to the least (8.1%) was including stories of successful recovery. The next lowest criteria adhered to by news media organizations was sharing resources (9.7%). The resource most frequently shared was the National Suicide

Prevention Line. The other two most frequently shared resources were the SAMHSA National Helpline and the Washington D.C. City Kids Wilderness Project.

Beyond individual-level interventions, 11 videos (17.7%) mentioned community-level factors and policies. For mental health and illness, examples included: city-level policy, the opioid crisis television show “Dopesick” and other multimedia created to raise awareness, federal legislation to address gun reform and student mental health, employment strikes to advocate for increased mental health support, and interventions led by community-based organizations.

Only 10 out of 62 videos (16.1%) mentioned suicide. Of those videos, only one conveyed that suicide is complex and often caused by multiple factors and only three conveyed that help is available. Two of the videos described the suicide method. The types of stories shared included: individuals dying by suicide (n=5), individuals having or not having suicidal ideation (n=3), national suicide rates increasing (n=1), and discussing the consequence of attacking a nuclear power plant as suicide (n=1). Of the five videos about individuals dying by suicide, two were military members and three were celebrities or public figures.

### Weather-related Disasters

There was a total of 276 weather-related disaster videos, but only 12 (4.3%) mentioned climate change. This suggests the connection between the frequency, intensity, and duration of weather-related disasters and climate change is not made clear by news media organizations. Table 5 shows the number and percentage of videos by news media organizations on weather-related disasters that met each Surgeon General recommendation from 2021. The 12 videos that mentioned climate change were removed from analysis, leaving 264 total weather-related disaster videos Examples of weather-related disasters covered during this time period include: Hurricane Ian in Florida, Kentucky tornadoes, Hurricane Fiona in Puerto Rico, California wildfires, extreme heat in the U.S. and Europe, and flash floods.

**Table 5.** Climate change videos that follow 2021 Surgeon General recommendations by news media organization.

| Surgeon General Recommendation | Total Videos n (%) | ABC News n (%) | CBS News n (%) | NBC News n (%) | Daily Wire n (%) |
| --- | --- | --- | --- | --- | --- |
| Total | 65 | 28 | 19 | 4 | 14 |
| No language that shocks, provokes, or creates a sense of panic | 33 (50.8%) | 17 (60.7%) | 12 (63.2%) | 1 (25.0%) | 3 (21.4%) |
| Entirely fact-based reporting | 35 (53.8%) | 25 (89.3%) | 8 (42.1%) | 2 (50.0%) | 0 (0.0%) |
| Ways public can make a positive difference | 7 (10.8%) | 4 (14.3%) | 3 (15.8%) | 0 (0.0%) | 0 (0.0%) |
| Positive Messages | 14 (21.5%) | 9 (32.1%) | 5 (26.3%) | 0 (0.0%) | 0 (0.0%) |
| Highlights actions to address the issue beyond the individual | 21 (32.3%) | 14 (50.0%) | 6 (31.6%) | 0 (0.0%) | 1 (7.1%) |
| Content warnings for distressing content<sup>a</sup> | 2 (8.0%)* | 1 of 9 (11.1%) | 0 of 10 (0.0%) | 1 of 4 (25.0%) | 0 of 2 (0.0%) |
| Average adherence rate<sup>b</sup> | 29.0% | 41.5% | 29.5% | 20.0% | 4.28% |

<sup>a</sup>25 out of 65 videos include distressing content. 2 out of 25 include a content warning.
<sup>b</sup>Only includes U.S. Surgeon General recommendations. Does not include the criteria added for the purposes of this study: actions beyond the individual.

The recommendation adherence rate ranged from 0.0% to 76.1% with media organizations least frequently following the recommendation to include ways the public can make a positive difference and most frequently following the recommendation to not include language that shocks, provokes, or creates a sense of panic. The criterion with the second highest adherence rate (75%) was to focus on entirely fact-based reporting. In combination, 155 (58.7%) weather-related disaster videos adhered to the top two recommendations, which on its own may seem positive. However, further analysis of the content showed a more complex picture. Many of these videos showed brief footage of the disaster unfolding or its aftermath with no reporting and only a simple caption or text on the video to explain the scene. While there were 155 videos that met the criteria of fact-based with no language that shocks, provokes, and creates a sense of panic, 135 videos (87.1%) failed to meet the criteria to exclude distressing content. Only 3 (2.4%) had a content warning.

The distressing nature of weather-related disasters is, in part, why the advisory recommends including positive messages and ways the public can make a positive difference. Only 33 videos (12.5%) included positive messages. Positive messages fell into the following categories: people or animal rescues; announcements of funding and other support for recovery from federal or state elected officials; and community members contributing to recovery efforts. Zero videos offered direct ways the viewing public can contribute to recovery efforts.

Only 22 videos (8.3%) highlighted actions to address the issue beyond the individual. Actions most frequently mentioned included government action to support recovery and emergency responders reporting to the scene. Only one video, from CBS News, mentioned proactive action to reduce future damage via a federal 10-year plan to “aggressively reduce catastrophic damage from wildfires.” Lastly, 220 (83.3%) included distressing content.

### Climate Change

A total of 65 videos mentioned “climate change,” “climate crisis,” or “global warming.” The recommendation adherence rate ranged from 8.0% to 53.8% with media organizations least frequently following the recommendation to include content warnings for distressing content and most frequently following the recommendation to focus on entirely fact-based reporting. Table 6 shows the number and percentage of videos by news media organizations on climate change that met each Surgeon General recommendation from 2021. The criterion with the second highest adherence rate (50.8%) was do not include language that shocks, provokes, or creates a sense of panic. Only 25 out of 65 videos (38.5%) included distressing content, and only two (8.0%) included a content warning.

**Table 6.**
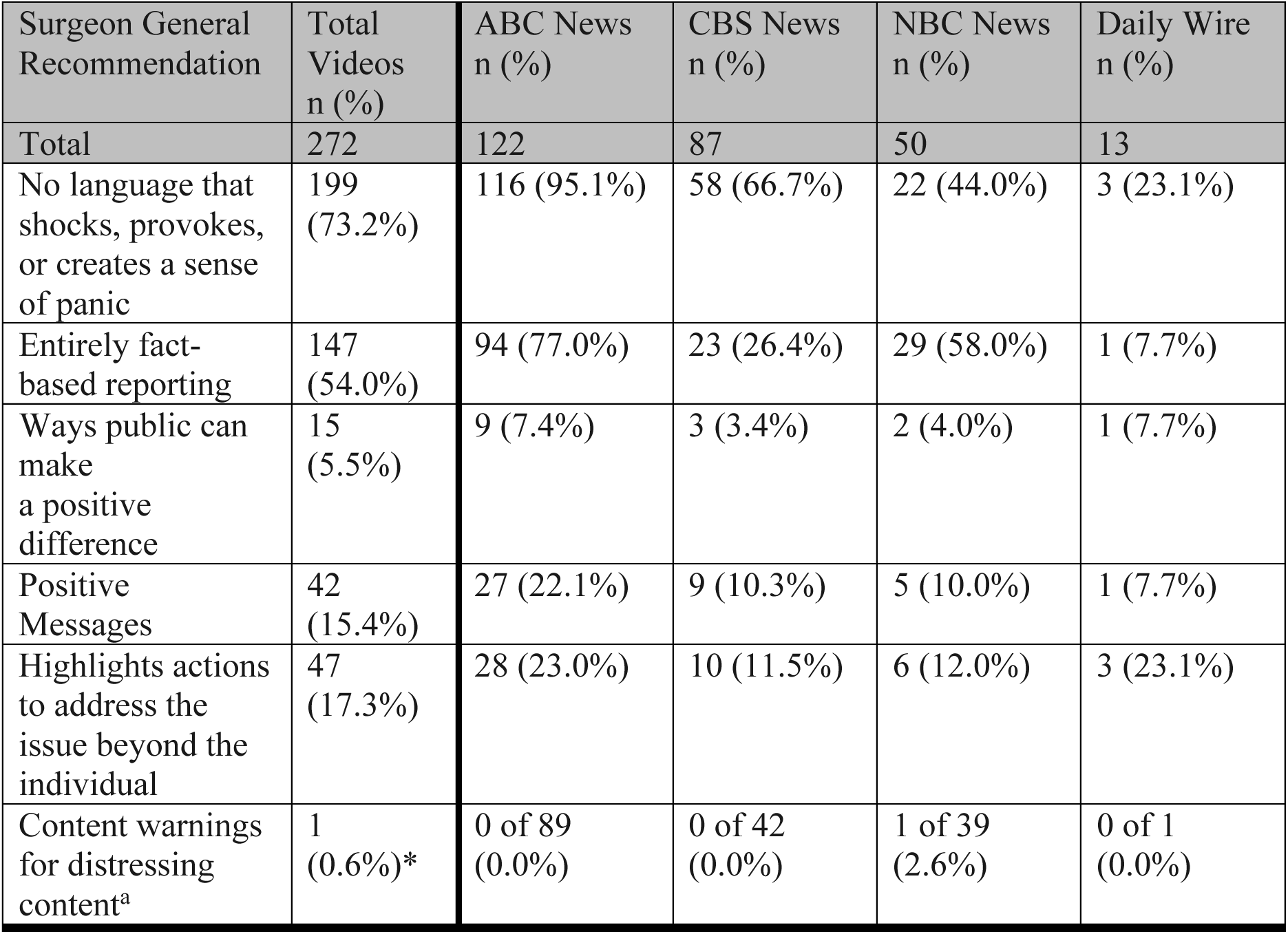
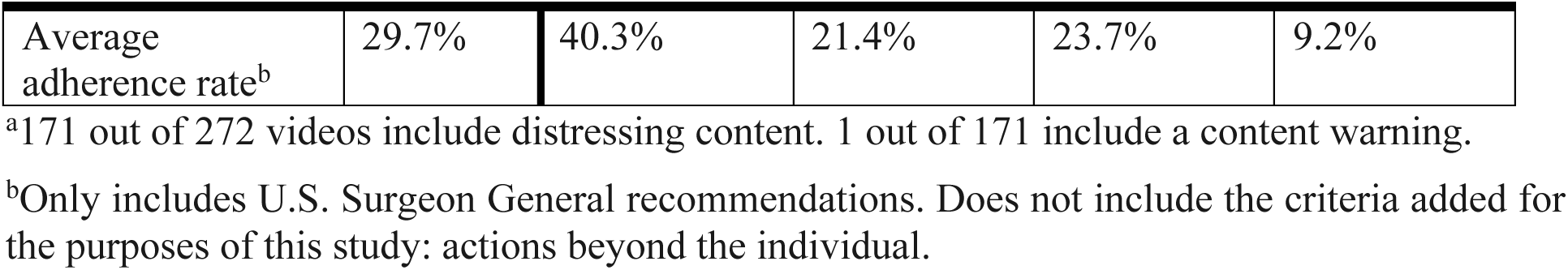
Mass shooting videos that follow 2021 Surgeon General recommendations by news media organization.

| Surgeon General Recommendation | Total Videos n (%) | ABC News n (%) | CBS News n (%) | NBC News n (%) | Daily Wire n (%) |
| --- | --- | --- | --- | --- | --- |
| Total | 272 | 122 | 87 | 50 | 13 |
| No language that shocks, provokes, or creates a sense of panic | 199 (73.2%) | 116 (95.1%) | 58 (66.7%) | 22 (44.0%) | 3 (23.1%) |
| Entirely fact-based reporting | 147 (54.0%) | 94 (77.0%) | 23 (26.4%) | 29 (58.0%) | 1 (7.7%) |
| Ways public can make a positive difference | 15 (5.5%) | 9 (7.4%) | 3 (3.4%) | 2 (4.0%) | 1 (7.7%) |
| Positive Messages | 42 (15.4%) | 27 (22.1%) | 9 (10.3%) | 5 (10.0%) | 1 (7.7%) |
| Highlights actions to address the issue beyond the individual | 47 (17.3%) | 28 (23.0%) | 10 (11.5%) | 6 (12.0%) | 3 (23.1%) |
| Content warnings for distressing content<sup>a</sup> | 1 (0.6%)* | 0 of 89 (0.0%) | 0 of 42 (0.0%) | 1 of 39 (2.6%) | 0 of 1 (0.0%) |
| Average adherence rate<sup>b</sup> | 29.7% | 40.3% | 21.4% | 23.7% | 9.2% |

<sup>a</sup>171 out of 272 videos include distressing content. 1 out of 171 include a content warning.
<sup>b</sup>Only includes U.S. Surgeon General recommendations. Does not include the criteria added for the purposes of this study: actions beyond the individual.

Roughly 1 out of every 2 climate change videos (50.8%) avoided language that shocks, provokes, or creates a sense of panic. The Daily Wire adhered to this recommendation the least (21.4%) while ABC News (60.7%) and CBS News (63.2%) adhered to this recommendation the most. Ways the public can make a positive difference were mentioned in 10.8% of videos and general positive messages were mentioned in 21.5% of videos. Videos mentioned consuming less, buying used clothing, planting trees, buying electric vehicles, and making your home more energy efficient as ways the public can make a positive difference. Positive messages fell into the following categories: stories about efforts to protect wildlife or the environment, mitigation or adaptation efforts to address climate change, and political or public figures (e.g. President Biden, King Charles, California Governor Newsom) taking action.

A higher percentage of videos (32.3%) mentioned actions to address climate change beyond the level of an individual. When compared to the other news organizations, ABC News adhered to this recommendation most frequently with half of their climate change videos mentioning actions beyond the individual. Examples include city-level action to address infrastructure resiliency to storms, state legislation such as banning carbon emitting vehicles in California, federal legislation such as the Inflation Reduction Act, and policy proposals that could better regulate plastics.

### Mass shootings

There was a total of 272 videos about mass shootings. Table 7 shows the number and percentage of mass shooting videos by news media organization that met each Surgeon General recommendation The recommendation adherence rate ranged from 0.6% to 73.2% with media organizations least frequently following the recommendation to include content warnings for distressing content and most frequently following the recommendation to not include language that shocks, provokes, or creates a sense of panic. ABC News had a higher adherence rate compared to other platforms for all U.S. Surgeon General criteria except for content warnings for distressing content. Most videos featured politicians or public figures, such as political commentators, celebrities, and police department officials, while some featured victims’ family members.

**Table 7.**
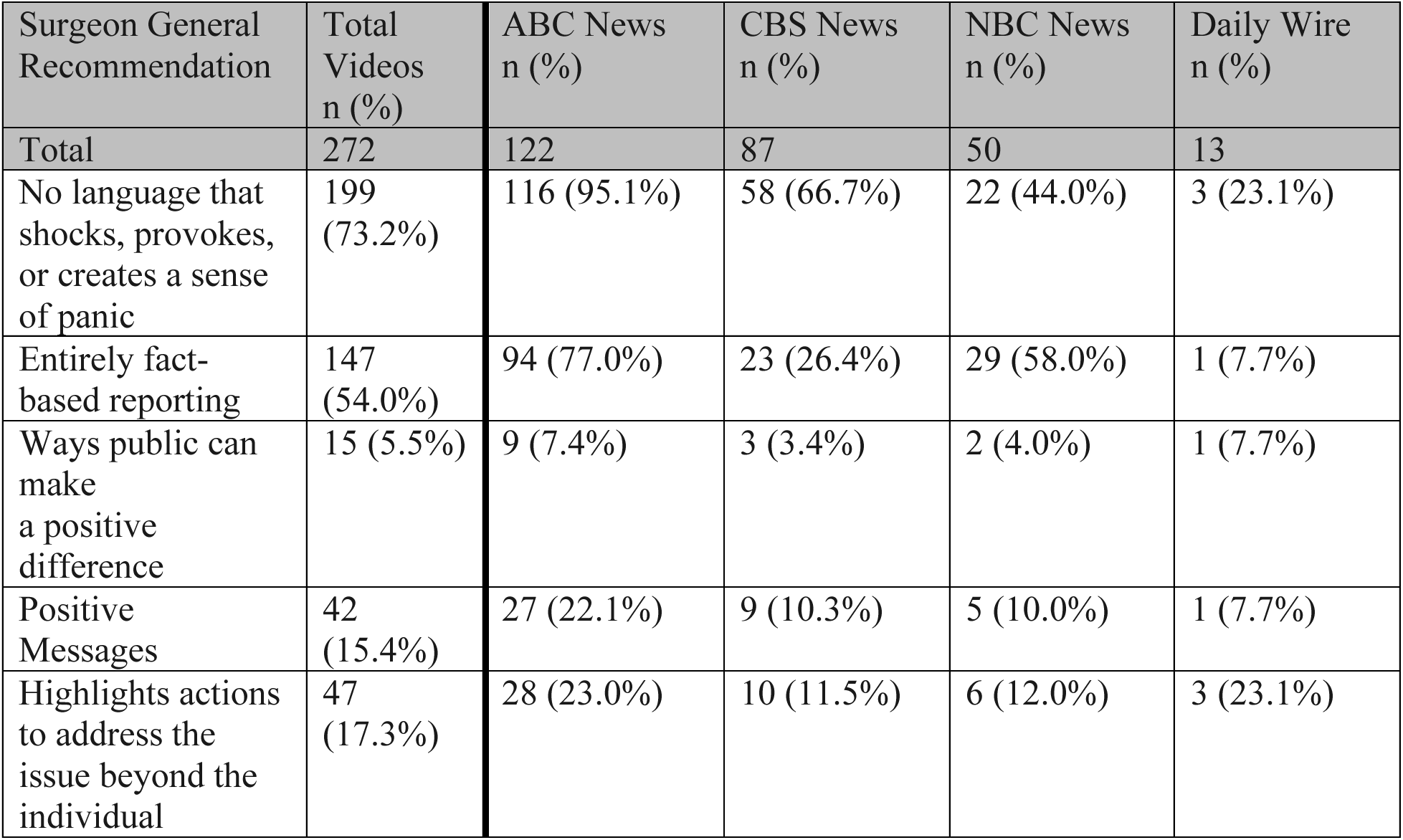
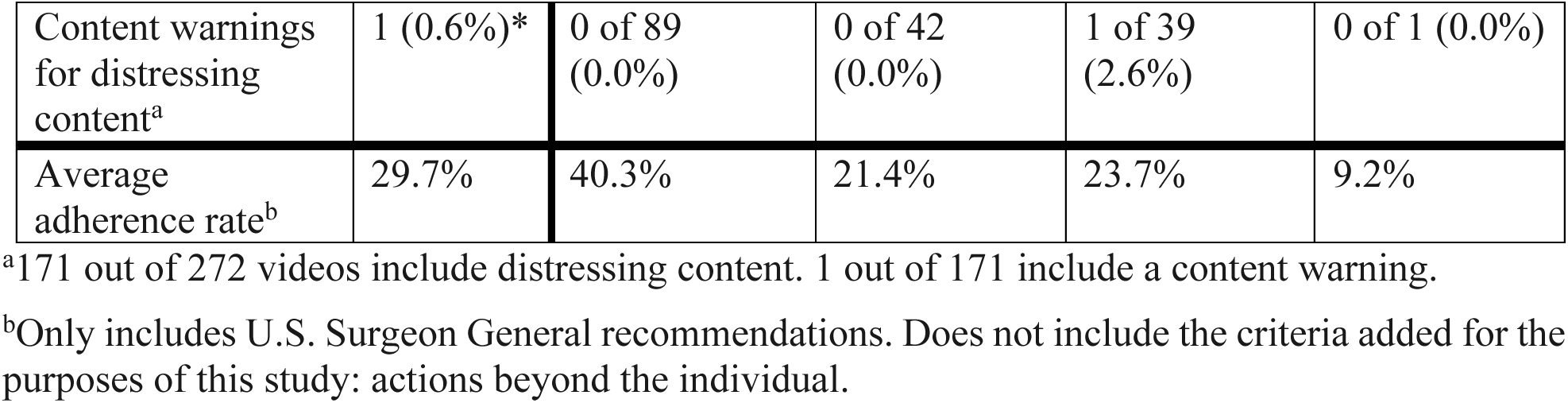
Mass shooting videos that follow Surgeon General recommendations by news media organization.

| Surgeon General Recommendation | Total Videos n (%) | ABC News n (%) | CBS News n (%) | NBC News n (%) | Daily Wire n (%) |
| --- | --- | --- | --- | --- | --- |
| Total | 272 | 122 | 87 | 50 | 13 |
| No language that shocks, provokes, or creates a sense of panic | 199 (73.2%) | 116 (95.1%) | 58 (66.7%) | 22 (44.0%) | 3 (23.1%) |
| Entirely fact-based reporting | 147 (54.0%) | 94 (77.0%) | 23 (26.4%) | 29 (58.0%) | 1 (7.7%) |
| Ways public can make a positive difference | 15 (5.5%) | 9 (7.4%) | 3 (3.4%) | 2 (4.0%) | 1 (7.7%) |
| Positive Messages | 42 (15.4%) | 27 (22.1%) | 9 (10.3%) | 5 (10.0%) | 1 (7.7%) |
| Highlights actions to address the issue beyond the individual | 47 (17.3%) | 28 (23.0%) | 10 (11.5%) | 6 (12.0%) | 3 (23.1%) |
| Content warnings for distressing content<sup>a</sup> | 1 (0.6%)* | 0 of 89 (0.0%) | 0 of 42 (0.0%) | 1 of 39 (2.6%) | 0 of 1 (0.0%) |
| Average adherence rate<sup>b</sup> | 29.7% | 40.3% | 21.4% | 23.7% | 9.2% |

<sup>a</sup>171 out of 272 videos include distressing content. 1 out of 171 include a content warning.
<sup>b</sup>Only includes U.S. Surgeon General recommendations. Does not include the criteria added for the purposes of this study: actions beyond the individual.

While 199 videos (73.2%) did not include language that shocks, provokes, or creates a sense of panic, the highest rate of adherence, videos that did conveyed children and communities are still in danger due to continued gun violence. The recommendation with the lowest adherence rate was to include content warnings for distressing content. Only one video out of 171 videos (0.01%) with distressing content included a warning.

Only 5.5% to 17.3% of content met recommendations to highlight positive messages, ways the public can make a difference, and actions to address the issue at a level beyond the individual. On June 25, 2022, President Biden signed the Bipartisan Safer Communities Act, known as the “first major gun safety legislation passed by Congress in nearly 30 years.”(25) Only three videos (3.4%) referenced this legislation passing both chambers and being signed into law.

Examples of positive messages shared included: describing possible policy solutions, communities coming together to pay tribute to the victims, and survivors expressing gratitude for being alive and witnessing people helping each other.

### COVID-19

There was a total of 305 videos about the COVID-19 pandemic. Table 8 shows the number and percentage of pandemic videos by news media organization that met each recommendation. The recommendation adherence rate ranged from 0.0% to 70.2% with media organizations least frequently following the recommendation to include content warnings for distressing content and most frequently following the recommendation to not include language that shocks, provokes, or creates a sense of panic.

**Table 8.** COVID-19 videos that follow 2021 Surgeon General recommendations by news media organization.

| Surgeon General Recommendation | Total Videos n (%) | ABC News n (%) | CBS News n (%) | NBC News n (%) | Daily Wire n (%) |
| --- | --- | --- | --- | --- | --- |
| Total | 305 | 109 | 86 | 21 | 89 |
| No language that shocks, provokes, or creates a sense of panic | 214 (70.2%) | 91 (83.5%) | 76 (88.4%) | 13 (61.9%) | 34 (38.2%) |
| Entirely fact-based reporting | 122 (40.0%) | 51 (46.8%) | 54 (62.8%) | 17 (81.0%) | 0 (0.0%) |
| Ways public can make a positive difference | 53 (17.4%) | 39 (35.8%) | 11 (12.8%) | 2 (9.5%) | 1 (1.1%) |
| Positive Messages | 109 (35.7%) | 58 (53.2%) | 33 (38.4%) | 11 (52.4%) | 7 (7.9%) |
| Highlights actions to address the issue beyond the individual | 73 (23.9%) | 47 (43.1%) | 18 (20.9%) | 4 (19.0%) | 4 (4.5%) |
| Content warnings for distressing content<sup>a</sup> | 0 (0.0%)* | 0 (0.0%) | 0 (0.0%) | 0 (0.0%) | 0 (0.0%) |
| Average adherence rate<sup>b</sup> | 32.7% | 43.9% | 40.5% | 41.0% | 2.25% |

<sup>a</sup>40 out of 301 videos include distressing content. 0 out of 40 include a content warning.
<sup>b</sup>Only includes U.S. Surgeon General recommendations. Does not include the criteria added for the purposes of this study: actions beyond the individual.

The Daily Wire adhered the least to the recommendation (38.2%) to not include language that shocks, provokes, or creates a sense of panic when compared to the other three news organizations, while CBS News (88.4%) and ABC News (83.5%) adhered the most. Most content not following this recommendation fell into three categories: (1) accusing Democrats of being a “liberal ruling class” implementing “full authoritarian tyranny” and setting up a “defense between everyday Americans and autocracy,” (2) claiming “the media don’t care about the truth” in pandemic reporting and (3) highlighting inconsistencies in implementation of regulations and calling COVID rules in “blue areas” “wildly stupid.” Shocking, provoking, or panic-inducing content from ABC, CBS, and NBC News was different, with language focused on the death toll, hospitals at “exploding capacities” unable to care for everyone, comparing the pandemic to a “war” and a “wildfire,” and the despair in not knowing if your loved one will survive and in believing “it’s hard to see an end in sight.”

To make a positive difference, news organizations highlighted getting vaccinated and masking. The Daily Wire, however, highlighted ways to resist pandemic regulations. Despite showing footage of body bags, strained hospitals, and an individual being forcefully taken into quarantine in China, no content warning was included on any of the 40 videos that included distressing content.

## Discussion

This study is the first to explore whether news media organization content on TikTok follows U.S. Surgeon General recommendations to protect youth mental health. Overall, these data show that news media organizations can significantly improve their adherence to federal recommendations. Given the link between stigmatizing or distressing media coverage and mental health (5), the observed lack of adherence to guidelines could have grave implications for youth mental health. This is particularly true given that the adherence rate was low for positive mental health and illness recommendations such as including stories of recovery and help-seeking, sharing resources, and using person-centered language compared to negative-exclusion criteria (ex. do not use stereotypes or demeaning language). While anti-stigma campaigns may have successfully discouraged the promotion of stereotypes and use of demeaning language, it is also important to move beyond avoiding stigma to promote positive messages about mental health.

For collective trauma content, the low adherence rates to recommendations are particularly concerning. Youth are grappling with existential threats and frequent media exposure related to extreme weather, mass shootings, and the pandemic. The coverage of these topics has the potential to exacerbate the youth mental health crisis if it focuses only – or mostly – on death tolls, worsening extreme weather events, strained hospital resources, and uncertainty instead of on positive messages and solutions.

One somewhat promising result regarding climate change content is that three out of four outlets did not call into question the existence of climate change, the anthropogenic cause of climate change, or the high level of alarm about the crisis. Daily Wire was the only outlet that called these elements into question in two videos. Of Daily Wire’s videos on climate change, a small number called into question the existence of global warming and its anthropogenic cause, while the remaining videos debated or criticized other points related to climate change. While the two videos including misinformation are concerning, the relatively small number of videos debating the existence and cause of climate change may indicate a shift in debate towards other aspects of the issue such as the appropriate level of alarm.

### Limitations and Future Research

There are limitations in both the data and analysis. The scope of the data included was limited. For example, the definition employed for mass shootings excluded media coverage of attempts thwarted by police, shootings with less than four fatalities such as the University of Virginia bus shooting, and shootings with no fatalities such as the New York City subway shooting on April 12, 2022 in which several people were injured. Future studies could use an expanded definition of mass shootings to understand a broader range of collective trauma events involving gun violence (e.g., Stanford’s Mass Shootings in America (26)). The study time frame consisted of one year to account for events that may occur at different times of the year; a longer time frame could give insight into trends across multiple years. Additionally, the content analyzed originated from only four outlets due to the size of the research team. While those four outlets have the most followers, including more outlets could give a better picture of the overall media landscape. Researchers may consider adding additional news outlets with more than 1 million followers on TikTok. Another comparison that future studies could investigate is legacy media performance versus emerging media performance – ABC, CBS, and NBC are legacy media organizations and performed better on average than Daily Wire; analyzing content from more emerging outlets could identify trends that are not exhibited by more established and traditional legacy media institutions. Lastly, with limited definitions in the advisory of “distressing content” and “language that does not shock, provoke, or create a sense of panic,” restrictive definitions were created and implemented for this study meaning data for these two criteria may be underestimates. Further clarity from the Surgeon General’s office can inform future research on measuring content that is distressing and includes shocking, provoking, panic-inducing language.

TikTok itself poses some limitations due to its proprietary algorithm and minimal data analytics are also limiting. For example, it is unknown to what extent youth were exposed to the videos included in this study. Without insight into how TikTok’s algorithm shares news media content with youth users, our data collection was undertaken under the assumption that news accounts with the most followers are more likely to be seen by users, many of whom are in the study’s age demographic. While TikTok content creators can see some follower data such as gender and where audiences are viewing from (27), these in-app metrics do not include age ranges and it is unclear if this data would even be accessible to third parties. Future studies could first verify a set of news media content that youth consumed and then analyze that data. Research involving TikTok may become easier as the company considers updates to its API for researchers (28).

### Practice-Based Recommendations

While behavioral interventions such as cognitive behavioral therapy or dialectical behavioral therapy could be used to encourage youth to moderate their consumption of distressing content, it is equally - if not more - important for interventions to target the content source: news media organizations. There are several recommendations for the U.S. Surgeon General’s office, advocacy organizations, and news media organizations to consider.

The advisory and its accompanying recommendations for whole-of-society efforts to address the youth mental health crisis is a critical first step. The Surgeon General’s office can go further by (1) adding additional recommendations, (2) clarifying certain definitions and criteria in their recommendations, and (3) promoting the recommendations more actively.

One recommendation to add for news media is: highlight actions that address the issue at a level beyond the individual. While the advisory does mention ways the public can make a positive difference, the examples shared are individual-led. Attention solely on individual actions could place unrealistic expectations on youth to solve problems that also require systemic solutions. Systemic actions such as state and federal policies are critical in addressing mental health, climate change, mass shootings, and other collective trauma events, and should receive adequate news coverage. The office should also encourage news media organizations to consider the purpose behind sharing distressing content knowing the impact it can have on youth mental health.

Secondly, the Surgeon General’s office should clarify two of their existing recommendations. First, a definition should be shared for language that “shocks, provokes, or creates a sense of panic.” While this study employed the aforementioned definition, it is important for news organizations to better understand what the Surgeon General means by this type of language so the recommendation is easier to follow. The office should also define distressing content with more specificity so news media can appropriately and consistently apply content warnings. Advocacy organizations (e.g., mental health coalitions, climate change organizations) should help promote these recommendations and educate news media organizations on best practices and incentivize staff to follow them. Social media and news media experts should also develop creative strategies to increase engagement on content that protects youth mental health.

## Conclusion

Public health research at the intersection of news and social media can help practitioners understand ways to address the youth mental health crisis. This study is the first to determine if news media organization content on TikTok is following U.S. Surgeon General recommendations to protect youth mental health. Low adherence rates to U.S. Surgeon General recommendations show that, overall, news media organizations’ content is not protecting youth mental health. Public health leaders can use these findings to inform interventions that ensure media organizations follow U.S. Surgeon General recommendations to protect youth and help end the mental health crisis. As the digital media landscape rapidly evolves and additional outlets enter the market, public health leaders will need to respond quickly and effectively to ensure news producers protect youth mental health.

## Data Availability

All data needed to evaluate the conclusions in the paper are present in the paper.

## Author contribution statement

All listed authors have contributed to the manuscript substantially and have agreed to the final submitted version. CH contributed to the conceptualization, methodology, data curation, investigation, formal analysis, writing of the original draft and reviewing and editing of writing. MP provided substantial reviewing and editing. DG, HS, VM, and RP contributed to formal analysis and reviewing and editing of writing. SL provided supervision and reviewing and editing of writing.

## Acknowledgement

Jonathan Purtle, PhD, for support as a Master’s thesis advisor.

## Notes

### Competing Interest Statement

The authors have declared no competing interest.

### Funding Statement

Yes

### Author Declarations

Ethics approval statement is not applicable, human subjects data was not analyzed.

